# SARS-Cov-2 RNA Found on Particulate Matter of Bergamo in Northern Italy: First Preliminary Evidence

**DOI:** 10.1101/2020.04.15.20065995

**Authors:** Leonardo Setti, Fabrizio Passarini, Gianluigi De Gennaro, Pierluigi Barbieri, Maria Grazia Perrone, Massimo Borelli, Jolanda Palmisani, Alessia Di Gilio, Valentina Torboli, Alberto Pallavicini, Maurizio Ruscio, Prisco Piscitelli, Alessandro Miani

## Abstract

In previous communications, we have hypothesized the possibility that SARS-CoV-2 virus could be present on particulate matter (PM) during the spreading of the infection, consistently with evidence already available for other viruses. Here, we present the first results of the analyses that we have performed on 34 PM10 samples of outdoor/airborne PM10 from an industrial site of Bergamo Province, collected with two different air samplers over a continuous 3-weeks period, from February 21st to March 13th. We can confirm to have reasonably demonstrated the presence of SARS-CoV-2 viral RNA by detecting highly specific RtDR gene on 8 filters in two parallel PCR analyses. This is the first preliminary evidence that SARS-CoV-2 RNA can be present on outdoor particulate matter, thus suggesting that, in conditions of atmospheric stability and high concentrations of PM, SARS-CoV-2 could create clusters with outdoor PM and, by reducing their diffusion coefficient, enhance the persistence of the virus in the atmosphere. Further confirmations of this preliminary evidence are ongoing, and should include real-time assessment about the vitality of the SARS-CoV-2 as well as its virulence when adsorbed on particulate matter. At the present, no assumptions can be made concerning the correlation between the presence of the virus on PM and COVID-19 outbreak progression. Other issues to be specifically addressed are the average concentrations of PM eventually required for a potential boost effect of the contagion (in case it is confirmed that PM might act as a carrier for the viral droplet nuclei), or even the theoretic possibility of immunization consequent to minimal dose exposures at lower thresholds of PM.

---

Severe acute respiratory syndrome known as COVID-19 disease - due to SARS-CoV-2 virus - is recognized to spread via respiratory droplets and close contacts.[1] The burden of COVID-19 was extremely severe in Lombardy and Po Valley (Northern Italy),[2] an area characterized by high concentrations of particulate matter, which are already known to have negative effects on human health.[3] Regional figures are available for Italy at the date of April 12^th^ show that about 30% of currently positive people still live in Lombardy (about 40% if considering the overall cases confirmed from the beginning of the epidemic), followed by Emilia Romagna (13.5% of currently positive people), Piedmont (10.5%), and Veneto (10%).[2] These four regions of the Po Valley account for 80% of total deaths recorded in Italy and 65% of Intensive Care Units admissions.[2] A research carried out by the Harvard School of Public Health seems to confirm an association between increases in PM concentrations and mortality rates due to COVID-19.[4]

In previous communications, we have hypothesized the possibility that SARS-CoV-2 virus could be present on particulate matter (PM) during the spreading of the infection, [5,6] consistently with evidence already available for other viruses [7-15]. However, the issue of airborne PM-associated microbiome, especially in urban environments, remains largely under-investigated [16], and – at the present – nobody has still carried out experimental studies specifically aimed at confirming or excluding the presence of the SARS-CoV-2 on PM.

Here, we present the first results of the analyses that we have performed on 34 PM10 samples of outdoor/airborne PM10 from an industrial site of Bergamo Province, collected with two air samplers (POS1 and POS2) over a continuous 3-weeks period, from February 21^st^ to March 13^th^.

Following the methodology described by Pan et al. in 2019 [17] for the collection, particle sizing and detection of airborne viruses, PM samples were collected on quartz fiber filters by using a low-volume gravimetric air sampler (38.3 l/min for 23 h), compliant with the reference method EN12341:2014 for PM10 monitoring. Particulate matter was trapped onto filters with 99.9% typical aerosol retention, properly stored and delivered to the laboratory of Applied and Comparative Genomics of Trieste University. Given the “environmental” nature of the sample, presumably rich in inhibitors of DNA polymerases, we proceeded with the extraction of RNA by using the Quick RNA fecal soil microbe kit adapted to the type of the filters.[18] Half filter was rolled, with the top side facing inward, in a 5 ml polypropylene tube, together with the beads provided in the kit. From the initial 1 ml of lysis buffer, we were able to get about 400 ul of solution, which was then processed as defined by the standard protocols, resulting in a final eluate of 15 ul. Subsequently, 5 ul were used for the SARS-CoV-2 testing. Given the particular origin of the sample, the qScript XLT 1-Step RT-qPCR ToughMix was used.[19] The amplification systems were those of the protocol developed by Corman et al, published on the WHO website [20].

The test was explicitly aimed at confirming or excluding the presence of the SARS-CoV-2 RNA on particulate matter. The first analysis used the “E gene” as a molecular marker and produced an impressive positive result on 15 out of 16 filters even if, as we could expect, the Ct was between 36-38 cycles. After that, we have replicated the analysis on 6 of the positive filters (already positive to “E gene”) by using the “RtDR gene” as a molecular marker – which is highly specific for SARS-CoV-2 – reaching 5 significant results of positivity; control tests to exclude false positivity were also successfully performed (Fig. 1).

**Fig.1.**
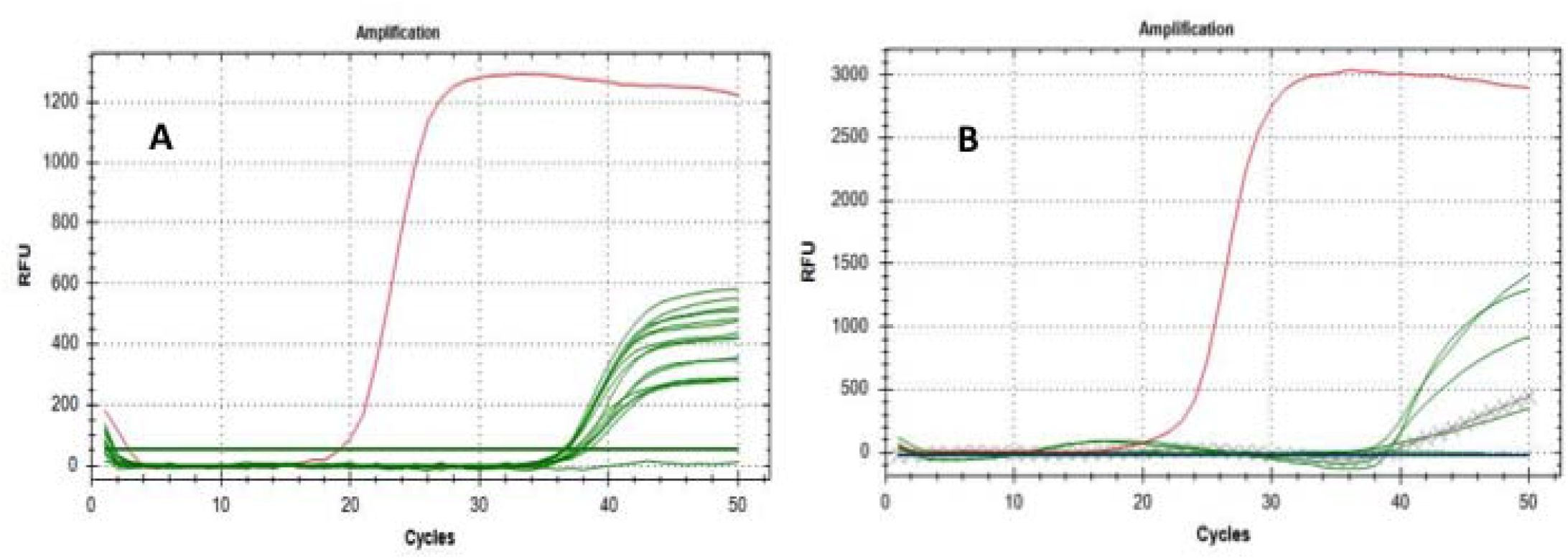
Amplification curves of E (A) and RdRP genes (B): green lines represent tested filters; cross line represents reference filter extractions; red lines represent the amplification of the positive samples.

**Fig.2.**
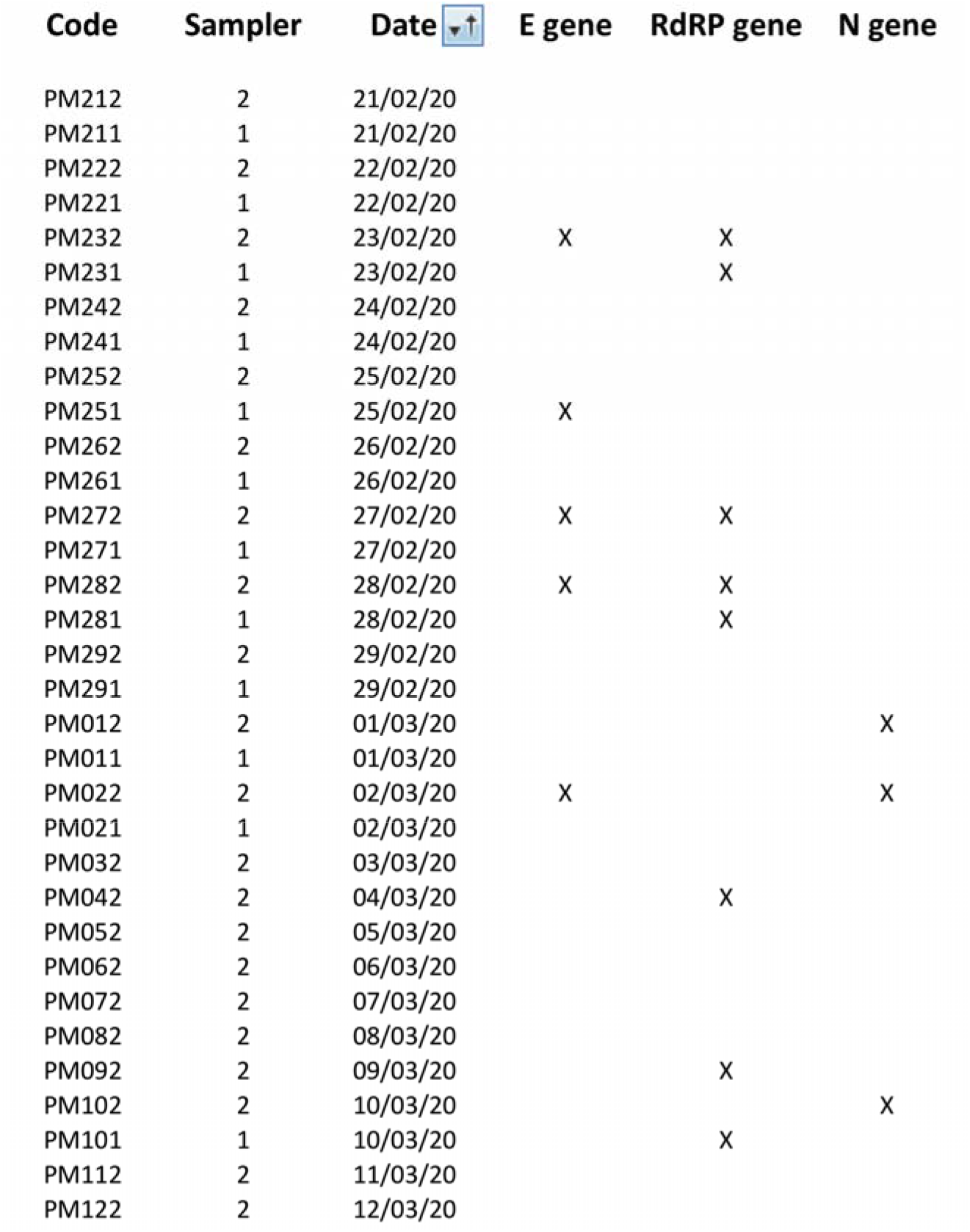
Positive results (marked with X) for E, N and RdRP genes obtained for all the PM10 samples tested by the two parallel genetic analyses.

To avoid the running out of the scarce sampling material available, the remaining extracted RNAs were delivered to the local University Hospital (one of the clinical centres authorized by the Italian Government for SARS-CoV-2 diagnostic tests), in order to perform a second parallel blind test. This second clinical laboratory tested 34 RNA extractions for the E, N and RdRP genes, reporting 7 positive results for at least one of the three marker genes, with positivity separately confirmed for all the three markers (Tab. 1). Because of the nature of the sample, and considering that the sampling has not been carried out for clinical diagnostic purposes but for environmental pollution tests (taking also into account that filters were stored for at least four weeks before undergoing molecular genetic analyses, as a consequence of the Italian shutdown), we can confirm to have reasonably demonstrated the presence of SARS-CoV-2 viral RNA by detecting highly specific “RtDR gene” on 8 filters. However, due to the lack of additional materials from the filters, we were not able to repeat enough number of tests to show positivity for all the 3 molecular markers simultaneously.

This is the first preliminary evidence that SARS-CoV-2 RNA can be present on outdoor particulate matter, thus suggesting that, in conditions of atmospheric stability and high concentrations of PM, SARS-CoV-2 could create clusters with outdoor PM and – by reducing their diffusion coefficient – enhance the persistence of the virus in the atmosphere. Further confirmations of this preliminary evidence are ongoing, and should include real-time assessment about the vitality of the SARS-CoV-2 as well as its virulence when adsorbed on particulate matter. At the present, no assumptions can be made concerning the presence of the virus on PM and COVID-19 outbreak progression. Other issues to be specifically addressed are the average concentrations of PM eventually required for a potential “boost effect” of the contagion in the areas experiencing the most dramatic burden of COVID-19, or even the theoretic possibility of immunization consequent to minimal dose exposures at lower thresholds of PM.

## Data Availability

Data available upon reasonable requests

## References

1. World Health Organisation, Modes of transmission of virus causing COVID-19: implications for IPC precaution recommendations, Scientific brief; available at: https://www.who.int/news-room/commentaries/detail/modes-of-transmission-of-virus-causing-covid-19-implications-for-ipc-precaution-recommendations (29 March 2020)

2. Italian Ministry of Health, daily bulletin Covid-19 outbreak in Italy, available at http://www.salute.gov.it/imgs/C_17_notizie_4451_0_file.pdf

3. EEA, European Environmental Agency, Air Quality in Europe 2019 Report; No 10/2019; European Environment Agency: Copenhagen, Denmark, availbale at: https://www.eea.europa.eu/publications/air-quality-in-europe-2019

4. Xiao Wu, Rachel C. Nethery, M. Benjamin Sabath, Danielle Braun, Francesca Dominici, Exposure to air pollution and COVID-19 mortality in the United States, available at: https://projects.iq.harvard.edu/files/covid-pm/files/pm_and_covid_mortality.pdf

5. Italian Society of Environmental Medicine (SIMA), Position Paper Particulate Matter and COVID-19, available at: http://www.simaonlus.it/wpsima/wp-content/uploads/2020/03/COVID_19_position-paper_ENG.pdf

6. Setti L., Passarini F., De Gennaro G., Barbieri P., Perrone M.G., Piazzalunga A., Borelli M., Palmisani J., Di Gilio A, Piscitelli P, Miani A., Is there a Plausible Role for Particulate Matter in the spreading of COVID-19 in Northern Italy?, BMJ Rapid Responses, April 8th 2020, available at: https://www.bmj.com/content/368/bmj.m1103/rapid-responses

7. Sedlmaier, N., Hoppenheidt, K., Krist, H., Lehmann, S., Lang, H., Buttner, M. Generation of avian influenza virus (AIV) contaminated fecal fine particulate matter (PM2.5): genome and infectivity detection and calculation of immission. Veterinary Microbiology. 139, 156–164 (2009)

8. Zhao, Y., Richardson, B., Takle, E., Chai, L., Schmitt, D., Win, H. Airborne transmission may have played a role in the spread of 2015 highly pathogenic avian influenza outbreaks in the United States. Sci Rep. 9, 11755. https://doi.org/10.1038/s41598-019-47788-z (2019)

9. Ma, Y., Zhou, J., Yang, S., Zhao, Y., Zheng, X. Assessment for the impact of dust events on measles incidence in western China. Atmospheric Environment. 157, 1–9 (2017)

10. Sorensen, J. H., Mackay, D. K. J., Jensen, C. Ø., Donaldson, A. I. An integrated model to predict the atmospheric spread of foot-and-mouth disease virus Epidemiol. Infect., 124, 577–590 (2000)

11. Glostera, J., Alexandersen, S. New Directions: Airborne Transmission of Foot-and-Mouth Disease Virus Atmospheric Environment, 38 (3), 503–505 (2004)

12. Reche, I., D’Orta, G., Mladenov, N., Winget, D.M., Suttle, C.A. Deposition rates of viruses and bacteria above the atmosperic boundary layer. The ISME Journal. 12, 1154–1162 (2018)

13. Qin, N., Liang, P., Wu, C., Wang, G., Xu, Q., Xiong, X., Wang, T., Zolfo, M., Segata, N., Qin, H., Knight, R., Gilbert, J.A., Zhu, T.F. Longitudinal survey of microbiome associated with particulate matter in a megacity. Genome Biology. 21, 55 (2020)

14. Zhao, Y., Richardson, B., Takle, E., Chai, L., Schmitt, D., Win, H. Airborne transmission may have played a role in the spread of 2015 highly pathogenic avian influenza outbreaks in the United States. Sci Rep. 9, 11755. https://doi.org/10.1038/s41598-019-47788-z (2019)

15. Ma, Y., Zhou, J., Yang, S., Zhao, Y., Zheng, X. Assessment for the impact of dust events on measles incidence in western China. Atmospheric Environment. 157, 1–9 (2017)

16. Jiang, W., Laing, P., Wang, B., Fang, J.,Lang, J., Tian, G., Jiang, J., Zhu, T.F. Optimized DNA extraction and metagenomic sequencing of airborne microbial communities. Nat. Protoc. 10, 768–779 (2015)

17. Pan, M., Lednicky, J.A., Wu, C.-Y., Collection, particle sizing and detection of airborne viruses. Journal of Applied Microbiology, 127, 1596-1611 (2019)

18. Zymoresearch Ldt, product description, available at: https://www.zymoresearch.com/products/quick-rna-fecal-soil-microbe-microprep-kit

19. Quantabio Ltd, descriprion of the product, available at: https://www.quantabio.com/qscript-xlt-1-step-rt-qpcr-toughmix

20. Corman, V. M., Landt, O., Kaiser, M., Molenkamp, R., Meijer, A., Chu, D. K., & Mulders, D. G. (2020) Detection of 2019 novel coronavirus (2019-nCoV) by real-time RT-PCR. Eurosurveillance, 25(3), available at:.https://www.ncbi.nlm.nih.gov/pmc/articles/PMC6988269/

